# From Breath to Brain: NICU Respiratory Interventions and Bedside Brain Signal Entropy Predict Later Autism Risk

**DOI:** 10.1101/2025.07.07.25331007

**Authors:** Madelyn G. Nance, Winnie R. Chang, Chad Aldridge, Jennifer Burnsed, Kevin Pelphrey, Santina Zanelli, Meghan H. Puglia

**Affiliations:** Department of Neurology, University of Virginia, Charlottesville, VA; Department of Pediatrics, Division of Neonatology, University of Virginia, Charlottesville, VA

**Keywords:** Prematurity, Ventilation, Autism Spectrum Disorder, Inflammation, EEG, Brain Signal Entropy

## Abstract

Premature infants often experience hypoxia and require prolonged ventilation, which can trigger systemic inflammation, damage the developing brain, and increase the risk of neurodevelopmental disorders such as Autism Spectrum Disorder (ASD). Early intervention is key for ensuring optimal outcomes for those with ASD; thus emphasizing the critical importance of accurately identifying infants at risk as early as possible. Here, infants underwent electroencephalography during social (held) and nonsocial (not held) resting state conditions to assess brain signal variability, saliva collection to determine inflammation, calculation of a novel Prognostic Respiratory Intensity Scoring Metric (PRISM) to assess the burden of respiratory support, and ASD testing in toddlerhood. Higher PRISM scores were associated with increased brain signal entropy during the nonsocial resting state. However, this association was not observed in the social resting state condition – particularly for male babies. Interestingly in female infants, we saw that the relationship between brain signal entropy and PRISM scores were potentially mediated by cytokines. Notably, the interaction between nonsocial resting state brain signal entropy, sex, and PRISM scores predicted risk of developing ASD with 88% accuracy. These non-invasive measures can identify infants at the highest risk for an ASD diagnosis before discharge.

## 1. Introduction

Most neural development occurs during the third trimester of pregnancy^1–3^. However, when this critical period of brain growth and maturation is interrupted by preterm birth before 37 weeks of gestation^4^, the typical processes underlying neurodevelopment are disrupted. In 2023, 10.3% of infants born in the United States were born preterm^5^. Thankfully, survival rates are continuously increasing^6^, an accomplishment which can be attributed to medical advancements made in the Neonatal Intensive Care Unit^7^ (NICU). The overwhelming majority of preterm infants hospitalized in the NICU will require some form of respiratory support during the neonatal period^8^. One of the most common consequences of prematurity is hypoxic injury^9^, which results from insufficient oxygen to support normal organ function^10^. Infants may experience 50 to 100 hypoxic events occurring a day^11^. To limit the consequences of hypoxia, physicians must use aggressive, lifesaving ventilation techniques^10^ that manually force large volumes of air into the lungs, compressing the heart and limiting blood flow to the brain^8^. As soon as fifteen minutes after the initiation of invasive mechanical ventilation, downstream damage to delicate, developing organs like the brain has begun^8,12^. While the exact mechanisms of ventilation-induced neonatal lung injury remain unclear, it has been documented that ventilation initiates an inflammatory response in which inflammatory cytokines, protein markers of inflammation, are elevated^13^. In preterm infants, the integrity of the blood-brain barrier is compromised following hypoxia^14^ and systemic inflammation^15^, ultimately leading to neurological and neurodevelopmental consequences.

Previous studies have identified the impact of early life, invasive respiration on developmental outcomes in toddlerhood. Walsh et. al. found that infants who experience protracted ventilation (for longer than 39 days) and assumed prolonged systemic inflammation are at an increased risk for impaired neurodevelopment as assessed by the Mental Development Index. Every infant included in the study who was ventilated for over 120 days had a neurodevelopmental impairment^16^. Similarly, Guillot et al. have observed that each additional day of ventilation predicts a decrease in motor scores as assessed by the Movement Assessment Battery for Children^17^. Specifically, preterm infants are at an increased risk of Autism Spectrum Disorder (ASD)^18^. The risk of being diagnosed with ASD is mediated by the degree of prematurity; there is a 1.4% chance of an infant born at term receiving an ASD diagnosis, as compared to a 6.1% chance if born extremely premature^18^. Male infants are at an abnormally high risk of receiving this diagnosis as there is a well established male bias in ASD prevalence^19^. Interestingly, male infants are also more likely to be born premature^20^, need more invasive forms of ventilation, develop chronic respiratory morbidities^17,21–24^, and experience adverse neurodevelopmental outcomes^23^. One study found that even when adjusting for gestational age, preterm male infants were more likely to die than preterm female infants^25^.

Here, we expand upon this previous work to identify associations between invasive early-life respiratory support, early markers of neurodevelopment, and later risk for autism in toddlerhood. To quantify early markers of neurodevelopment, we use electroencephalography (EEG), a non-invasive method for measuring electrical brain activity that can be conducted safely at the bedside. Brain signal variability is increasingly investigated in the context of neurodevelopment, as it plays an important role in facilitating synchronous neural firing and the development of neural networks that underlie later cognitive function^26–31^. In the present study, we leverage EEG to quantify multiscale entropy – a measure of brain signal variability that assesses the random and deterministic properties of neural signals across multiple time scales^32^. Shorter time scales are thought to reflect neural processing within local networks, whereas longer time scales capture processing in more widespread cortical networks^33,34^. Given the vulnerability of male infants to the effects of prematurity^17,21–24^, we hypothesized that they would exhibit significant differences in brain signal entropy across varying levels of respiratory support, even when held by a caregiver. We postulated that this relationship between early life respiratory support and entropy would be mediated by an infant’s inflammatory profile specifically in male infants, and that brain signal entropy, sex, and an infant’s respiratory experience in the NICU could accurately predict autism risk in toddlerhood.

## 2. Methods

### 2.1. Participants

EEG data was collected from 158 preterm infants hospitalized and recruited from the University of Virginia NICU between 0-4 months of age as part of a larger, ongoing neurodevelopmental study. Parental consent was obtained for all participants. After preprocessing 65 infants had sufficient artifact-free EEG data. Of those 65 infants, 34 had saliva samples collected for protein analysis at the time of EEG, and 17 returned to the lab at a corrected age of 24 months for autism testing. Infant demographics for the 65 infants with usable EEG data are detailed in Table 1. For a total breakdown of participant overlap, see Figure S1 in the Supplement.

**Table 1:** Participant Demographics for the 65 infants that had usable EEG data.

| Degree of Prematurity |  |
| --- | --- |
| GA (weeks), mean, SD | 31.2, 3.5 |
| Extremely Preterm (<28 weeks) | 16 (25%) |
| Very Preterm (28-32 weeks) | 11 (17%) |
| Moderate Preterm (32-34 weeks) | 26 (40%) |
| Late preterm (34-37 weeks) | 12 (18%) |
| PMA at EEG Collection |  |
| GA (weeks) mean, SD | 37.1, 3.2 |
| **Multiplicity** |  |
| Singleton | 45 (69%) |
| Twin | 18 (28%) |
| Triplet | 2 (3%) |
| **Sex** |  |
| Female | 32 (49%) |
| Male | 33 (51%) |
| **Race** |  |
| White | 49 (75%) |
| Black | 11 (17%) |
| More than 1 Race | 2 (3%) |
| Other | 3 (4%) |
| **Respiratory Diagnoses** |  |
| None | 11 (17%) |
| Respiratory Distress Syndrome | 35 (54%) |
| Chronic Lung Disease | 19 (29%) |
| **Neonatal Medical Complications** |  |
| Retinopathy of Prematurity | 25 (38%) |
| Intraventricular Hemorrhage | 10 (15%) |

### 2.2. EEG Acquisition and Analysis

Full scalp resting state EEG was recorded using 32 Ag/AgCl active actiCAP slim electrodes (Brain Produces GmBh, Germany) affixed to an elastic cap in the 10-20 electrode placement. EEG was amplified with a BrainAMP DC Amplifier and digitized using BrainVision Recorder software with a sampling rate of 5000 Hz, online referenced to FCz, and online band-pass filtered between 0.1 and 1000 Hz. Each infant included in the study participated in both a social resting state, where they rested in a caregiver’s arms, and a nonsocial resting state, where the infant lay in their bassinet. The order of these conditions was randomized across participants. Each condition lasted for a maximum of 7 minutes or until the infant was no longer compliant.

Data preprocessing and entropy calculation was accomplished with the Automated Preprocessing Pipe-Line for the Estimation of Scale-wise Entropy from EEG Data (APPLESEED)^35^ implemented with EEGLab v2022.1^36^ in MATLAB v2020a. The resampling rate was set to 500 Hz, the data were then band-pass filtered from 0.1-100 Hz and separated into 10,000 ms epochs. On average, each infant had 42.02 epochs for the social resting state condition and 39.31 epochs for the nonsocial resting state condition. Epochs contaminated with extreme artifacts (>500 µV) were removed, and data underwent independent component analysis.

Problematic channels were identified and removed using the adjusted_ADJUST algorithm^37^. Subjects with more than 30% of channels removed were rejected. On average, 4.42 components were removed from the social resting state condition and 4.55 components were removed from the nonsocial resting state condition. Epochs in which the standard deviation exceeded 80 µV within a 200-ms sliding window with a 100-ms window step were discarded as artifacts, and channels identified as problematic via the FASTER algorithm^38^ were interpolated. Subjects with more than 35% of channels interpolated were rejected. On average, 3.25 channels were interpolated from the social resting state condition and 4.0 channels were interpolated from the nonsocial resting state condition. Participants with fewer than 3 artifact free trials after preprocessing were removed. Clean EEG data was then referenced to the average of all scalp electrodes. Due to the high attrition rate observed in pediatric EEG data^39^, three epochs from the social resting state and three from the nonsocial resting state with a total global field power (GFP) closest to the median GFP for each participant were selected for inclusion in the entropy calculation^29^. Selecting these trials helps to minimize the risk of trials in the dataset that are contaminated with high residual amplitude artifacts^29^. Multiscale (scale-wise) entropy was computed on these trials using APPLESEED^35^ with a similarity criterion r = 0.5, and a pattern length m = 2. Entropy values were averaged across scales and channels to create mean entropy values for the social and nonsocial resting state conditions.

### 2.3. Saliva Collection and Analysis

We collected passive saliva from each infant in the NICU using a small plastic pipette on the day of EEG collection. Immediately following collection, saliva was stored in microcentrifuge tubes frozen at −30 degrees Celsius before being transferred to a −80 degree Celsius freezer at the UVA Flow Cytometry Core Facility. Flow cytometry is a cell separation technique utilizing the unique ways in which proteins refract light and fluorescence to identify the cellular contents of a sample^40^. Prior to analysis samples were thawed and 50 uL of assay buffer was added. Cytokines were then fluorescently labeled with fluorochromes using a Human Antibody Th17 panel from Millipore Sigma and run in duplicate on a Luminex MAGPIX platform which uses an ELISA-derived immunoassay. Cytokines produced by Th17 cells are crucial for immune responses at mucosal sites such as the trachea and lungs^41^. Th17 immune responses are elevated in individuals with autism^42^ and there is evidence that Th17 cells are expressed in neonatal cells^43^. This panel generated concentrations of 24 measured cytokines (IL-17F, GM-CSF, IFN-γ, IL-10, MIP-3α, IL-12p70, IL-13, IL-15, IL-17A, IL-22, IL-9, IL-1β, IL-33, IL-2, IL-21, IL-4, IL-23, IL-5, IL-6, IL-27, IL-31, TNF-α, TNF-β, and IL-28a) and further analysis utilized analyte concentrations based on the average of replicate samples.

### 2.4. Autism Diagnostic Testing

At 24 months’ corrected age (M= 21.1 months corrected, SD= 1.7), participants returned for autism testing. The Autism Diagnostic Observation Schedule-2nd edition (ADOS-2)^44^ is a gold-standard, semi-standardized, direct observation autism assessment. It consists of a series of structured, observed activities that allow a research-reliable examiner to observe social interactions, communication skills, creativity, play, and restricted and repetitive behaviors. Here, the ADOS-2 Toddler module (used for children 30 months or younger) was administered. Scores for this module are calculated to yield a Range of Concern (Little-to-No-Concern, Mild-to-Moderate Concern, and Moderate-to-Severe Concern) rather than autism cutoffs^44^.

### 2.5. PRISM Score Calculation

PRISM scores are a summative indicator of respiratory health in the NICU that accurately quantify the invasive nature of respiratory support. They are used here to help advance our understanding of how respiratory needs impact long term neurodevelopmental outcomes. To calculate PRISM for each of the 65 infants included in analysis, we identified the most invasive resuscitation method used in the delivery room and each delivery room resuscitation type was assigned a score We next used custom R^45^ scripts to extract the most frequently charted ventilation type on each day of an infant’s hospital stay and summed the consecutive days the infant received that form of ventilation to determine its duration. Ventilation types were categorized based on the invasiveness of the respiratory support, assigned a score, and then multiplied by the episode’s duration to generate a ventilation score. For each infant, the ventilation score was summed and then added to the delivery room score to generate a final PRISM score. PRISM score calculation is described in detail in Nance et. al^46^.

We assessed the distribution of PRISM scores among included subjects and visualized a rightward skew with several high yet clinically meaningful values. To avoid inappropriately impacting the relationship between PRISM scores and other variables, we split PRISM scores into quartile categories. Infants in the highest quartile were classified as critical, followed by severe (third quartile), moderate (second quartile), and mild (lowest quartile).

### 2.6. Statistical Analysis

We first investigated the effects of resting state condition, PRISM score category, and sex on an infant’s brain signal entropy. To do so, we conducted a repeated measures ANOVA on the 65 subjects with usable brain signal entropy data and PRISM scores using the within subjects factors of condition (social vs. nonsocial resting state) and scale (0.15 - 250 Hz) as well as the between subject factors of PRISM score category (critical, severe, moderate, mild) and sex (male, female). We also conducted a post-hoc Tukey’s Honestly Significant Differences (HSD) test within each combination of sex and condition and adjusted for multiple comparisons with Bonferroni correction. Additionally, we investigated the impact of region of interest (Frontal, Central, Parietal, Occipital, and Temporal), resting state condition, and PRISM score category on an infant’s brain signal entropy. The results of this analysis can be seen in the Supplement.

To determine how an infant’s sex and inflammatory profile mediates the relationship between their PRISM scores and brain signal entropy across resting state conditions, we conducted Multi-Group Partial Least Squares - Path Modelling (MG-PLS-PM) using the “plspm” package in R^45^. PLS-PM is a predictive modeling approach used to study the complex and multivariate associations between measured and latent variables^47^. PSL-PM is optimal for use with smaller sample sizes than are required for other multivariate modelling techniques^47,48^. We defined three latent constructs: PRISM scores and brain signal entropy which were modeled as single-indicator latent variables, and cytokines which were modeled as a reflective construct composed of 24 cytokines. The value of each cytokine was increased by one and log transformed^49–51^. Three subjects had seven missing cytokine values due to low concentration and were excluded from analysis. One subject with one missing cytokine value was retained and the value imputed with the mean of that specific cytokine’s values across other participants. The PLS-PM algorithm was implemented on each combination of infant sex and condition and standardized latent variable scores estimated through iterative weight adjustment. Model performance was evaluated with path coefficients, standard errors, R^2^ values, and Goodness-of-Fit.

To determine our ability to use early-life biological markers to predict later autism risk, we fit two multinomial logistic regression models to predict later ADOS Range of Concern from sex, PRISM scores, and brain signal entropy in the nonsocial and social resting state. We assessed model performance by calculating the accuracy (fraction of correct classifications over total classifications), precision (ratio of true positives to all identified positives), recall (sensitivity / correct identification of a true positive) and F1 score (measure of accuracy from precision and recall) for each ASD risk class. To visualize model performance and demonstrate the balance between sensitivity and specificity across various thresholds, we conducted a receiver operating characteristic (ROC) analysis for each model using the pROC^52^ package in R^45^. We constructed one-vs-all binary outcomes for each multi-class outcome (Little-to-No Concern, Mild-to-Moderate Concern, and Moderate-to-Severe Concern) as well as the micro-average and macro-average ROC curves. For each ROC curve, we calculated the area under the curve (AUC) values with a 95% Confidence Interval (CI) using a 10,000 iteration bootstrap resampling technique.

## 3. Results

### 3.1. When male infants with the highest PRISM scores are held by a caregiver, their brain signal variability is indistinguishable from their healthier counterparts

We ran a repeated measures ANOVA to determine the impact of sex, resting state condition (social vs. nonsocial), PRISM category, and scale on brain signal entropy (Figure 1). There was a significant interaction effect for both condition x PRISM category and PRISM category x sex. To further investigate these interactions, we ran Tukey’s HSD tests within each combination of sex and condition, the results of which can be seen in Table 2.

**Figure 1.**
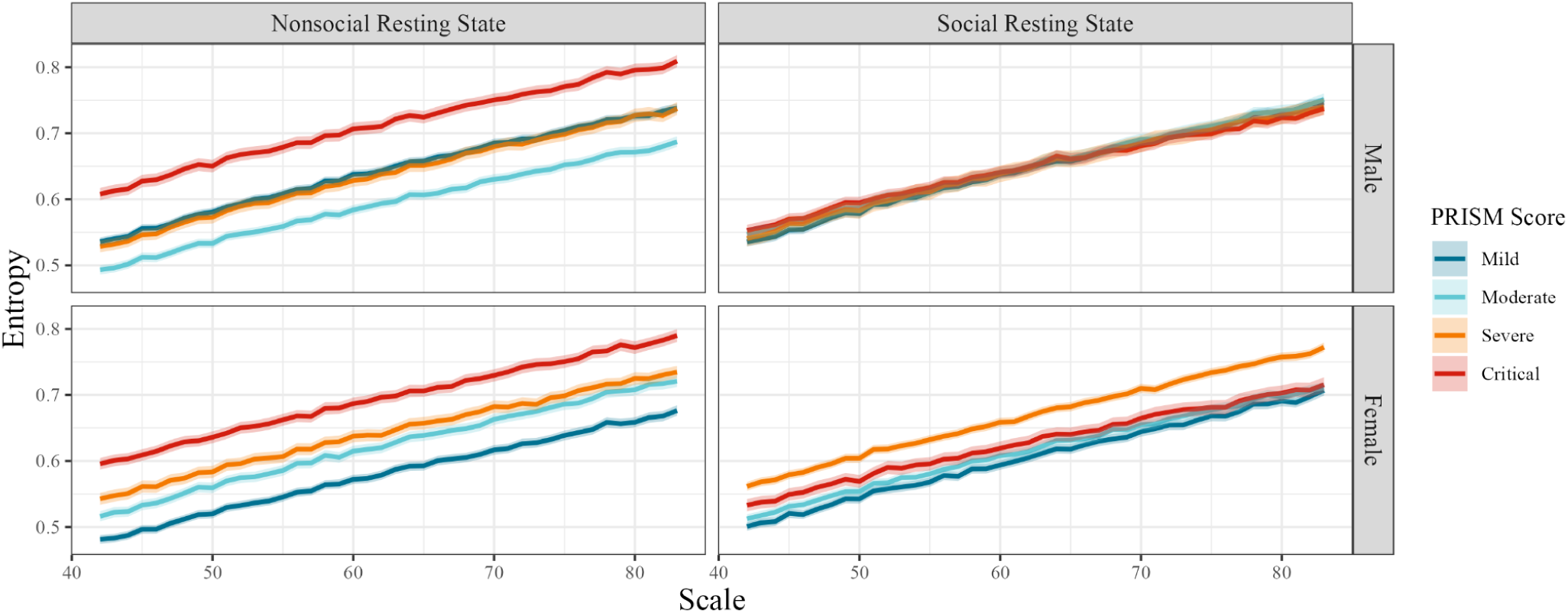
**When male infants with the highest PRISM scores are held by a caregiver, their brain signal variability is indistinguishable from their healthier counterparts.**Within the nonsocial resting state condition, both female and male infants exhibit significant differences in entropy between the majority of PRISM categories. In the social resting state condition, female infants retain significant differences in entropy between several PRISM categories. However, for male babies in the social resting state condition, differences in entropy converge and there are no significant differences in entropy between any PRISM categories ( p = 1.00 for all).

**Table 2:** The results of the repeated measures ANOVA and post-hoc Tukey’s HSD investigating differences in entropy between PRISM categories across resting state conditions and Sex.

| Effect | Degrees of Freedom | F statistic | P value |
| --- | --- | --- | --- |
| Condition | 1 | 1.22 | 0.269 |
| PRISM Category | 3 | 86.09 | < 0.001 *** |
| Scale | 1 | 1944.82 | < 0.001 *** |
| Sex | 1 | 33.10 | < 0.001 *** |
| Condition x PRISM Category | 3 | 61.30 | < 0.001 *** |
| Condition x Scale | 1 | 0.01 | 0.940 |
| PRISM Category x Scale | 3 | 0.64 | 0.586 |
| Condition x Sex | 1 | 1.29 | 0.255 |
| PRISM Category x Sex | 3 | 28.68 | < 0.001 *** |
| Scale x Sex | 1 | 0.06 | 0.804 |
| Condition x PRISM Category x Scale | 3 | 0.32 | 0.812 |
| Condition x PRISM Category x Sex | 3 | 13.84 | < 0.001 *** |
| Condition x Scale x Sex | 1 | 0.04 | 0.837 |
| PRISM Category x Scale x Sex | 3 | 0.11 | 0.952 |
| Condition x PRISM Category x Scale x Sex | 3 | 0.27 | 0.850 |
| **Comparison** | **Mean Difference** | **95% Confidence Interval** | **Adjusted P value** |
| **Females** |  |  |  |
| **Nonsocial Resting State** |  |  |  |
| Severe vs. Critical | -0.05 | [-0.08, -0.03] | < 0.001 *** |
| Moderate vs. Critical | -0.07 | [-0.10, -0.05] | < 0.001 *** |
| Mild vs. Critical | -0.11 | [-0.14, -0.09] | < 0.001 *** |
| Moderate vs. Severe | -0.02 | [-0.04, 0.00] | 0.230 |
| Mild vs. Severe | -0.06 | [-0.09, -0.04] | < 0.001 *** |
| Mild vs. Moderate | -0.04 | [-0.07, -0.02] | < 0.001 *** |
| **Social Resting State** |  |  |  |
| Severe vs. Critical | 0.04 | [0.01, 0.07] | < 0.001 *** |
| Moderate vs. Critical | -0.01 | [-0.04, 0.01] | 0.858 |
| Mild vs. Critical | -0.02 | [-0.05, 0.00] | 0.136 |
| Moderate vs. Severe | -0.05 | [-0.08, -0.03] | < 0.001 *** |
| Mild vs. Severe | -0.06 | [-0.09, -0.04] | < 0.001 *** |
| Mild vs. Moderate | -0.01 | [-0.04, 0.01] | 0.866 |
| **Males** |  |  |  |
| **Nonsocial Resting State** |  |  |  |
| Severe vs. Critical | -0.08 | [-0.10, -0.05] | < 0.001 *** |
| Moderate vs. Critical | -0.12 | [-0.15, -0.09] | < 0.001 *** |
| Mild vs. Critical | -0.07 | [-0.10, -0.04] | < 0.001 *** |
| Moderate vs. Severe | -0.04 | [-0.07, -0.02] | < 0.001 *** |
| Mild vs. Severe | 0.01 | [-0.02, 0.03] | 0.998 |
| Mild vs. Moderate | 0.05 | [0.02, 0.08] | < 0.001 *** |
| **Social Resting State** |  |  |  |
| Severe vs. Critical | -0.002 | [-0.03, 0.03] | 1.000 |
| Moderate vs. Critical | 0.002 | [-0.03, 0.03] | 1.000 |
| Mild vs. Critical | -0.003 | [-0.03, 0.02] | 1.000 |
| Moderate vs. Severe | 0.004 | [-0.02, 0.03] | 1.000 |
| Mild vs. Severe | -0.001 | [-0.03, 0.03] | 1.000 |
| Mild vs. Moderate | -0.005 | [-0.03, 0.02] | 1.000 |

### 3.2. Cytokines mediate the pathway between PRISM scores and brain signal entropy in female infants

We conducted MG-PLS-PM to determine how an infant’s inflammatory profile mediates the relationship between their PRISM scores and brain signal entropy in both male and female infants in the nonsocial and social resting state (Figure 2). We ensured that each cytokine indicator was loaded properly onto the Cytokines construct by ensuring that the outer loading of each indicator was above 0.7^53^. The only cytokine indicator that did not meet this threshold was MIP3A. There were no significant relationships between any of the constructs for either sex in the social resting state condition (Table 3). In the social resting state condition, model fit was higher for female infants (Goodness-of-Fit = 0.34) than male infants ( Goodness-of-Fit =0.22). The model utilizing female infants explained 25% of the variance in brain signal entropy (R^2^) and the model utilizing male infants explained 19% of the variance. For male infants in the nonsocial resting state condition, PRISM scores were significantly positively associated with brain signal entropy. However for female infants in the nonsocial resting state, cytokines have a trending positive association with brain signal entropy (Table 3). In the nonsocial resting state condition, model fit was higher for male infants (Goodness of Fit = 0.48) than female infants (Goodness of Fit =0.37). The model using female infants explained 31% of the variance in brain signal entropy (R^2^) and the model using male infants explained 45% of the variance.

**Figure 2.**
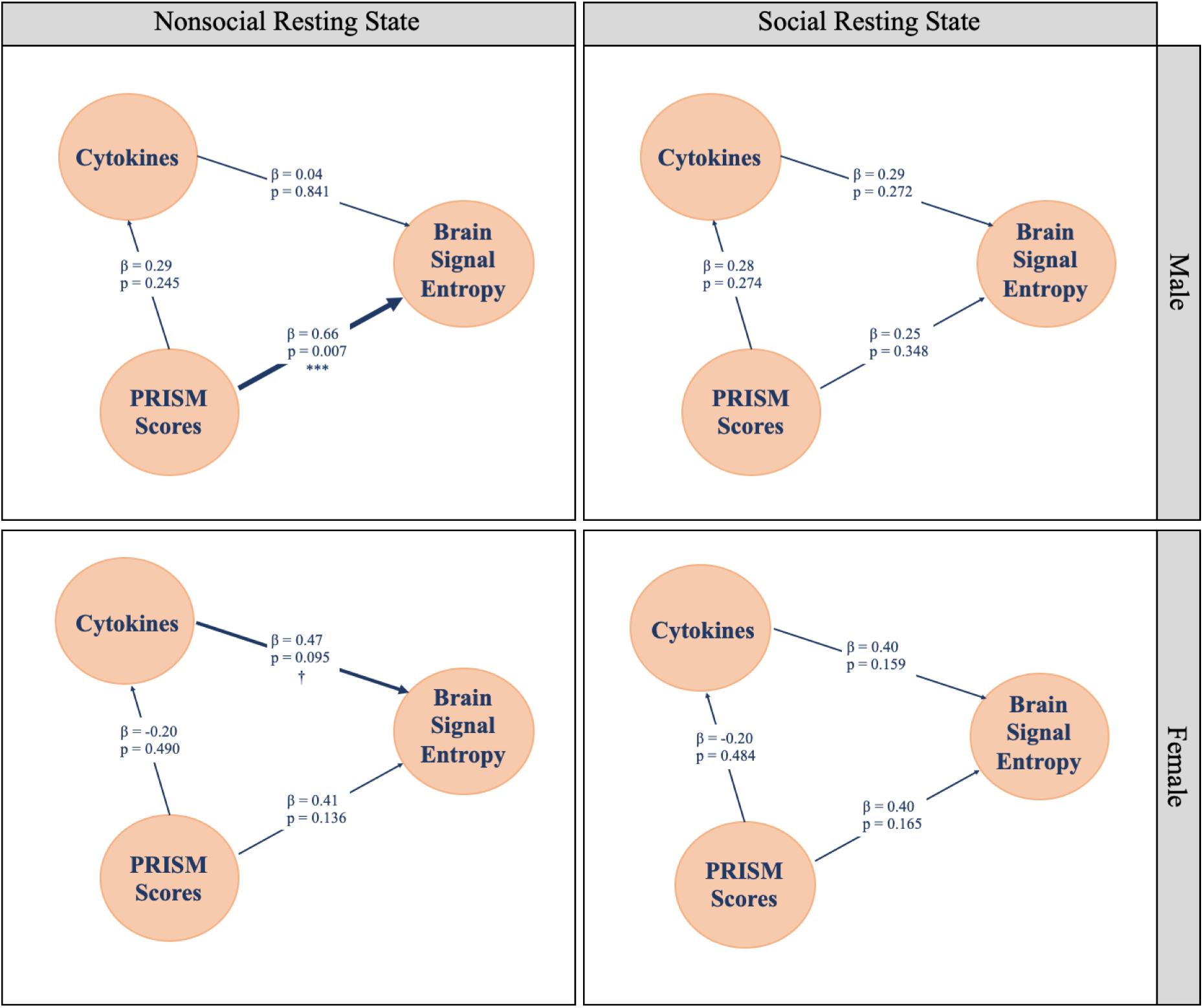
Cytokines mediate the pathway between PRISM scores and brain signal entropy in female infants. There are no significant constructs significantly associated with brain signal entropy in the social resting state condition for either sex. In the nonsocial resting state condition, the construct significantly associated with brain signal entropy differs based on sex. PRISM scores are positively associated with brain signal entropy in males while cytokines have a trending positive relationship with brain signal entropy in females.

**Table 3:** Path coefficients, (Standard Errors), and p values for a PLSPM modelling the relationship between PRISM scores, cytokines, and brain signal entropy.

|  |  |  | From: |  |
| --- | --- | --- | --- | --- |
|  |  | To: | PRISM Scores | Cytokines |
| Nonsocial Resting State | Male | Brain Signal Entropy | 0.66 (0.21)<br>p = 0.007 *** | 0.04 (0.21)<br>p = 0.841 |
|  |  | Cytokines | 0.29 (0.25)<br>p = 0.245 | - |
|  | Female | Brain Signal Entropy | 0.41 (0.26)<br>p = 0.136 | 0.47 (0.26)<br>p = 0.095 † |
|  |  | Cytokines | -0.20 (0.28)<br>p = 0.490 | - |
| Social Resting State | Male | Brain Signal Entropy | 0.25 (0.25)<br>p = 0.348 | 0.29 (0.25)<br>p = 0.272 |
|  |  | Cytokines | 0.28 (0.25)<br>p = 0.274 | - |
|  | Female | Brain Signal Entropy | 0.40 (0.27)<br>p = 0.165 | 0.40 (0.27)<br>p = 0.159 |
|  |  | Cytokines | -0.20 (0.28) | - |
| | | | $p = 0.484$ | |

### 3.3. The interaction between brain signal entropy, PRISM score, and sex accurately predicts autism risk

In an exploratory analysis, we ran two multinomial logistic regressions using sex, PRISM score, and social or nonsocial resting state brain signal entropy to predict ADOS Range of Concern. The model using brain signal entropy from the nonsocial resting state condition predicted the infant’s ADOS Range of Concern with an overall accuracy of 88% (Figure 3).

**Figure 3.**
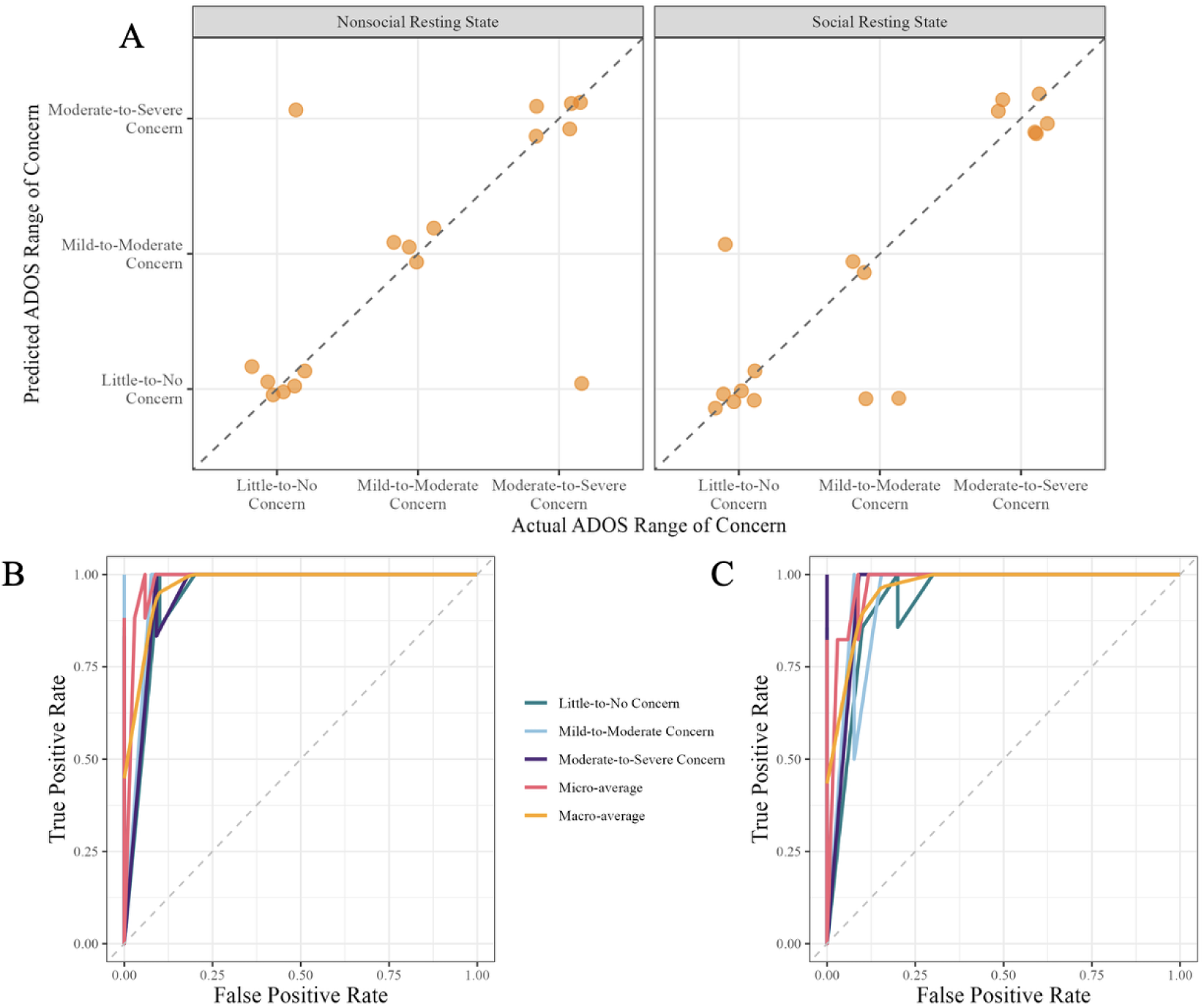
The interaction between brain signal entropy, PRISM score, and sex accurately predicts autism risk with 88% accuracy. **A.** A multinomial logistic regression model was trained using an infant’s PRISM score, brain signal entropy (during social or nonsocial resting state condition), and sex to predict their ADOS Range of Concern. A scatter plot of predicted and actual diagnosis visualizes the 88% accuracy of the model when using the nonsocial resting state condition and the 82% accuracy of the model when using the nonsocial resting state condition. **B.** The performance of the multinomial logistic regression model trained using the nonsocial resting state brain signal entropy, sex, and PRISM scores to predict ADOS Range of Concern as well as the micro and macro-average curves were visualized with an ROC analysis. The AUCs for all of these classes are included in Table 4. **C.** The performance of the multinomial logistic regression model trained using the social resting state brain signal entropy, sex, and PRISM scores to predict ADOS Range of Concern as well as the micro and macro-average curves were visualized with an ROC analysis. The AUCs for all of these classes are included in Table 4.

**Table 4:**
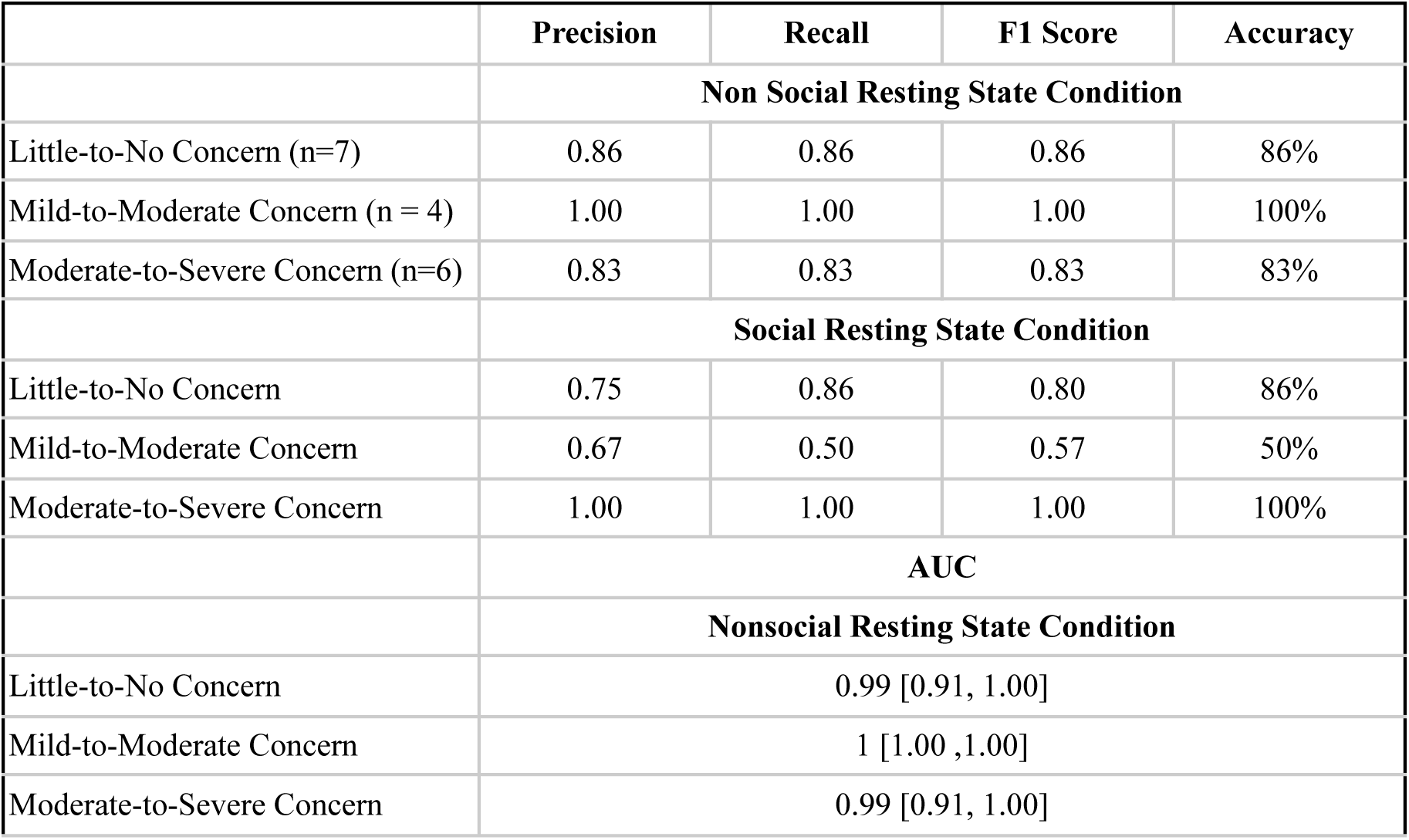
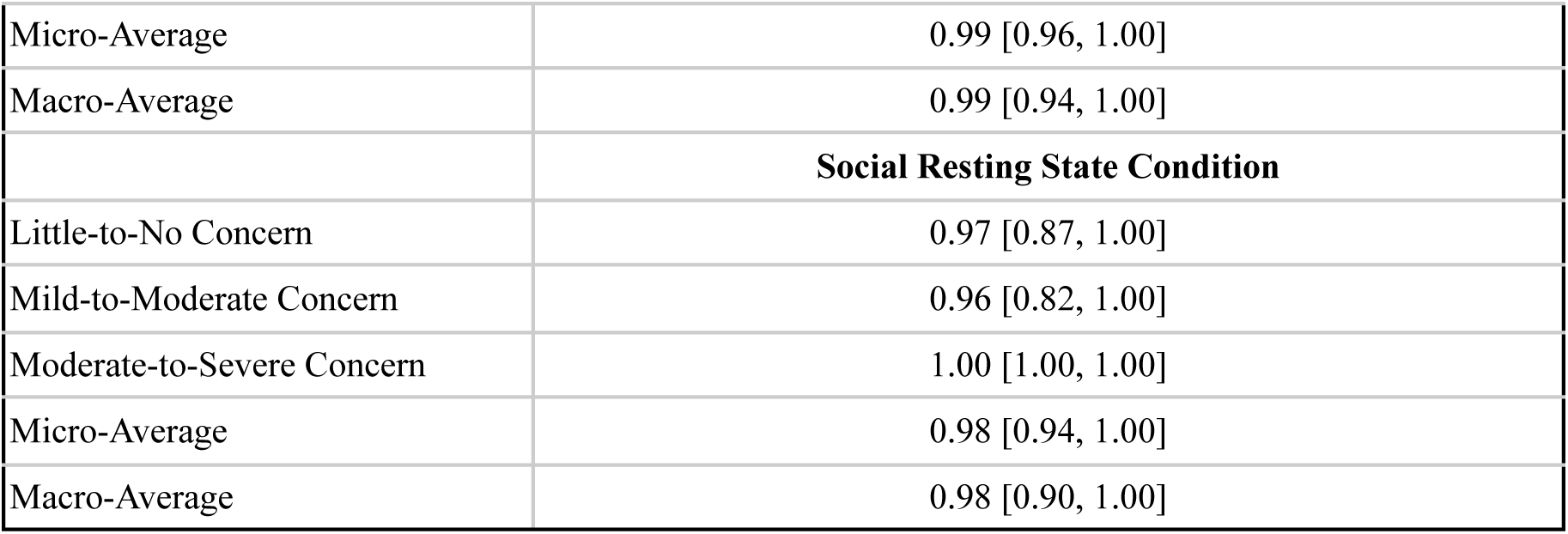
Results of 2 multinomial logistic regression models predicting ADOS Range of Concern from PRISM scores, sex, and brain signal entropy from the social and nonsocial resting state conditions respectively. The table also includes the results of a ROC analysis assessing both model’s classification performance.

|  | Precision | Recall | F1 Score | Accuracy |
| --- | --- | --- | --- | --- |
|  | **Non Social Resting State Condition** |  |  |  |
| Little-to-No Concern (n=7) | 0.86 | 0.86 | 0.86 | 86% |
| Mild-to-Moderate Concern (n = 4) | 1.00 | 1.00 | 1.00 | 100% |
| Moderate-to-Severe Concern (n=6) | 0.83 | 0.83 | 0.83 | 83% |
|  | **Social Resting State Condition** |  |  |  |
| Little-to-No Concern | 0.75 | 0.86 | 0.80 | 86% |
| Mild-to-Moderate Concern | 0.67 | 0.50 | 0.57 | 50% |
| Moderate-to-Severe Concern | 1.00 | 1.00 | 1.00 | 100% |
|  | **AUC** |  |  |  |
|  | **Nonsocial Resting State Condition** |  |  |  |
| Little-to-No Concern | 0.99 [0.91, 1.00] |  |  |  |
| Mild-to-Moderate Concern | 1 [1.00 ,1.00] |  |  |  |
| Moderate-to-Severe Concern | 0.99 [0.91, 1.00] |  |  |  |

**Table 4:** Results of 2 multinomial logistic regression models predicting ADOS Range of Concern from PRISM scores, sex, and brain signal entropy from the social and nonsocial resting state conditions respectively.
|  |  |
| --- | --- |
| Micro-Average | 0.99 [0.96, 1.00] |
| Macro-Average | 0.99 [0.94, 1.00] |
| **Social Resting State Condition** |  |
| Little-to-No Concern | 0.97 [0.87, 1.00] |
| Mild-to-Moderate Concern | 0.96 [0.82, 1.00] |
| Moderate-to-Severe Concern | 1.00 [1.00, 1.00] |
| Micro-Average | 0.98 [0.94, 1.00] |
| Macro-Average | 0.98 [0.90, 1.00] |

Per-class performance metrics and the AUC for each class can be seen in Table 3. For this model, the AUC for each class was exceptionally high and well above the 0.75 clinical threshold for utility.

When using brain signal entropy recorded during the social resting state condition, the model predicted ADOS Range of Concern with an overall accuracy of 82%. It is of particular interest that this model only performed at chance (50% accuracy) when predicting which infants would have a Mild-to-Moderate ADOS Range of Concern. Per-class performance metrics and the AUC for each class can be seen in Table 4.

## 4. Discussion

It is critically important that we identify infants who experience prolonged, invasive respiratory support and are at risk for neuroinflammation as early as possible, given that this inflammation may disrupt neural development and increase the risk of later neurodevelopmental disorders^54,55^. The predictive models presented here represent the first integrated effort to incorporate respiratory support needs into outcome prediction in preterm infants. To capture an infant’s neural functioning we used brain signal entropy derived from non-invasive, bedside EEG recordings collected during resting state. Resting state EEG provides valuable information into the organization of early life cortical rhythms and provides a window into baseline brain activity^43^ that can be predictive of later cognitive function^56^, and has been shown to differ between preterm and term infants^56^, making it a valuable tool for assessing neural maturation and development in this population. In this paper, we recorded resting state EEG during two distinct conditions: in the social resting state condition, infants were held by a caregiver and during the nonsocial resting state condition, the infants lay in their bassinets. By utilizing two resting state conditions, we attempt to capture differences in social touch experienced by infants in the NICU. Despite the well established fact that simple touch interventions are beneficial to infants’ health^57–60^ and later cognitive development^61–64^, premature infants commonly miss out on positive forms of touch^65^. This lack of touch is only more common in the most critically ill infants who, one study found, were held by their parents only twice a week on average^65^.

Here we sought to understand how the complex interactions that exist between an infant’s early life respiratory experiences, inflammatory states, neural function, and later developmental outcome differ by sex. We hypothesized that male infants would exhibit significant differences in brain signal entropy across varying levels of respiratory support, even when held by a caregiver. When we examined the effects of sex and resting state conditions on differences in brain signal entropy across PRISM categories (critical, severe, moderate, and mild), we identified a significant three way interaction between resting state condition, PRISM category, and sex. In the nonosical resting state, across both sexes, brain signal entropy varies significantly across PRISM categories. Specifically, within the nonsocial resting state, infants with critical PRISM scores have the highest levels of entropy and the infants with mild / moderate scores the lowest. In the social resting state condition, differences in brain signal entropy between PRISM categories in female infants remain significant. However, when male infants are held by their caregiver, these differences completely converge. These findings suggest that male infants may be more sensitive to the beneficial impacts of simple social touch interventions such as kangaroo care or neonatal massage techniques.

Although the mechanisms linking early invasive respiratory interventions to later neurodevelopmental disorders are not fully understood, one possible explanation is that hypoxic events, which require respiratory support, trigger systemic inflammation^13^. Hypoxia-induced brain injuries often manifest as white matter damage because pre-oligodendrocytes (Pre-OLs), the myelinating cells of the cortex^66^, are highly vulnerable to hypoxia^67^. During a hypoxic event, the brain is deprived of the oxygen energy needed to operate the intracellular pumps that regulate ion balance across the neuronal membrane^10^. This leads to high levels of intracellular calcium, which triggers the release of toxic amounts of glutamate into the synaptic cleft^68,69^. Persistent activation of glutamate receptors by calcium ultimately results in excitotoxicity and the death of the neuron^68,69^. Pre-OLs are especially susceptible to this process because they express a high density of calcium receptors^68^. As many as 32.8% of premature infants have white matter damage when assessed by Magnetic Resonance Imaging (MRI)^70^. Recent research using bedside EEG has demonstrated potential in both identifying hypoxic injury^71^ and utilizing EEG to predict adverse MRI outcomes and neurodevelopmental outcomes at two years^72^. While further studies should directly examine the impact of post-hypoxic inflammation on white matter using MRI, we can determine how systemic inflammation interacts with brain signal entropy, a metric shown to be associated with synchronous neuronal firing and the development of neural networks underlying later cognitive function^26–31^, two functions that can also be linked to white matter^73^.

We postulated that this relationship between early life respiratory support and entropy would be mediated by an infant’s inflammatory profile specifically in male infants. To examine this, we conducted MG-PLS-PM to explore the relationships that exist in our sample between the need for early life respiratory support, inflammation as assessed by inflammatory cytokines, and brain signal entropy. Within the nonsocial resting state condition we observed differences in the significant predictor of brain signal entropy based on sex. For male infants, PRISM scores were a significant positive predictor of brain signal entropy. However, among female infants, PRISM scores were not significantly associated with brain entropy and instead, cytokine levels showed a trending positive association with brain signal entropy. This is contrary to our hypothesis however, exploratory research has shown that female infants have a more active immune system^74^ that matures with gestation^75^ as compared to their male counterparts. Therefore, our results may reflect the immaturity of the male immune system in early life rather than a true lack of inflammatory impact on neural functioning. Within the social resting state condition, we saw no significant associations between PRISM scores and inflammation on brain signal entropy. Overall, models in the social resting state condition had lower goodness-of-fit and R^2^ values than those in the nonsocial condition. The social regions of the brain, and the white matter connections between them, are some of the last to develop and are likely to be interrupted by preterm birth^76^. The absence of significant associations between brain signal entropy while infants are held, their PRISM scores, and cytokine levels may reflect the immature neural systems not yet capable of processing or responding to the salience of a social context. However, it is important to consider that the sample size for this analysis is small and future studies should explore these associations within larger groups.

We next hypothesized that the interaction between sex, PRISM category, and brain signal entropy in the social or nonsocial resting state condition could be used to predict the risk of ASD in toddlerhood. We fit two multinomial logistic regression models using sex and PRISM category and either social or nonsocial resting state brain signal entropy. Overall, the model using entropy from the nonsocial resting state had a higher accuracy (88%) than the one using entropy from the social resting state condition (82%). While the success of nonsocial resting state brain signal entropy may be surprising, given that ASD often manifests itself as deficits in social interaction^77^, in recent years it has becoming increasingly clear that nonsocial resting state paradigms can reveal information about spontaneous neural activity and neural synchronization that are altered in autistic individuals^78^. Perhaps early in life, this intrinsic neural organization is more predictive of a later diagnosis than early interactions with social stimuli. Given the clinical applicability of this predictive model for the early detection of ASD we conducted an ROC analysis and calculated the AUC for each binary outcome. AUCs are widely used for assessing the diagnostic accuracy of clinical models and an AUC above 0.9 is considered excellent while 1.0 indicates perfect discernment^79^. Our model had AUCs for each outcome that are considered excellent and discernment of Mild-to-Moderate risk for ASD was perfect, demonstrating this model has the potential to be implemented in clinical settings.

Many preterm infants receive an ASD diagnosis later in life despite the fact that early diagnosis and intervention is critical to better outcomes^80^. As many as 50% of infants who are born at an extremely low birth weight miss subsequent neurosensory screenings that are essential for identifying those at risk of developing ASD. Additionally, one study found that only 53% of high risk infants returned for a follow up visit between 18-36 months^81^, a crucial window of neural plasticity where early interventions may be maximally effective^82^. The process of receiving a diagnosis and initiating the appropriate intervention is complicated by many factors including systemic barriers to developmental care. Black and hispanic mothers are less likely to be referred to follow up programs^83^, and receiving a diagnosis is significantly hindered by having public insurance or living in a rural area^84^. Additionally, preterm infants are less likely to have consistent family medicine providers^85^ who are the main practitioners of premature follow-up care and initiate necessary developmental testing^86^.

Here, we present a predictive model including only factors that can be determined *before* discharge, a critical window after which many families are lost to follow up. A family’s race, education status, and income should not influence their infant’s neurodevelopmental outcomes and neither should the ventilators capable of saving an infant’s life. The interactions between PRISM scores, brain signal entropy, and sex can help to identify infants at a high risk for ASD, allowing their parents to be armed with the information that they need to advocate and secure support for their child before they leave the hospital, regardless of their discharge destination.

### 4.1. Limitations and Future Directions

The sample size of this study did not allow for the incorporation of the concentration of inflammatory cytokines into the predictive model for risk of ASD. Additionally, small sample sizes increase the risk for overfitting in multinomial logistic regression models and as such in future research, these models should be fit to larger sample sizes^87^ to ensure their validity.

The model presented in this manuscript was tested only at the University of Virginia’s level IV NICU. Future studies should adopt a multisite approach to assess the generalizability of the model’s predictive performance across hospitals and neonatal populations. Additionally, given the compelling impact of social touch on brain signal entropy in our sickest infants, future studies should incorporate simple holding interventions to determine whether increased physical contact alleviates differences in brain signal entropy across PRISM categories.

### 4.2. Conclusion

Premature infants have underdeveloped lungs, experience frequent hypoxic events and require prolonged respiratory support. Invasive mechanical ventilation, when prolonged, is associated with systemic inflammation which may adversely impact brain development and increase the risk for neurodevelopmental impairments. These delays are one of the most common consequences of prematurity, and it is crucial that we identify infants at an increased risk as early as possible. However, these diagnoses are difficult to obtain, expensive, and influenced by systemic biases that stymie many parent’s’ ability to obtain appropriate interventions for their children. By building a model that takes into account infant sex, objective and non-invasive measures of brain signal variability, and the summation of respiratory interventions experienced in the NICU, we can identify the infants who are at the highest risk of an ASD diagnosis before they ever leave the hospital.

## Data Availability

All data produced in the present study are available upon reasonable request to the authors

## Acknowledgements

The cytometric data for this manuscript were generated in the University of Virginia Flow Cytometry Core Facility (RRid:SCR_017829) and is partially supported by the NCI Grant (P30-CA044579). We would like to thank the UVA STAR (Supporting Transformative Autism Research) team for conducting the autism assessments included in this study. Their expertise and dedication were instrumental to this work. Finally, we would like to thank each and every infant family who participated in this research.

## 4.3. CRediT authorship contribution statement

**Madelyn G. Nance -** Conceptualization, Writing - original draft, Writing-Review and Editing, Formal Analysis, Methodology, Investigation. **Winnie R. Chang -** Project administration, Writing-Review and Editing. **Chad Aldridge -** Data Curation, Writing-Review and Editing**. Jennifer Burnsed-**Writing-Review and Editing , **Kevin Pelphrey -** Writing-Review and Editing, **Santina Zanelli -**Writing-Review and Editing , **Meghan H. Puglia -** Conceptualization, Funding Acquisition, Supervision, Writing-Review and Editing

## 4.4. Data Statement

Data will be made available upon reasonable request.

