## Supplementary Materials for "From Breath to Brain: NICU Respiratory Interventions and Bedside Brain Signal Entropy Predict Later Autism Risk"

### **Supplement**

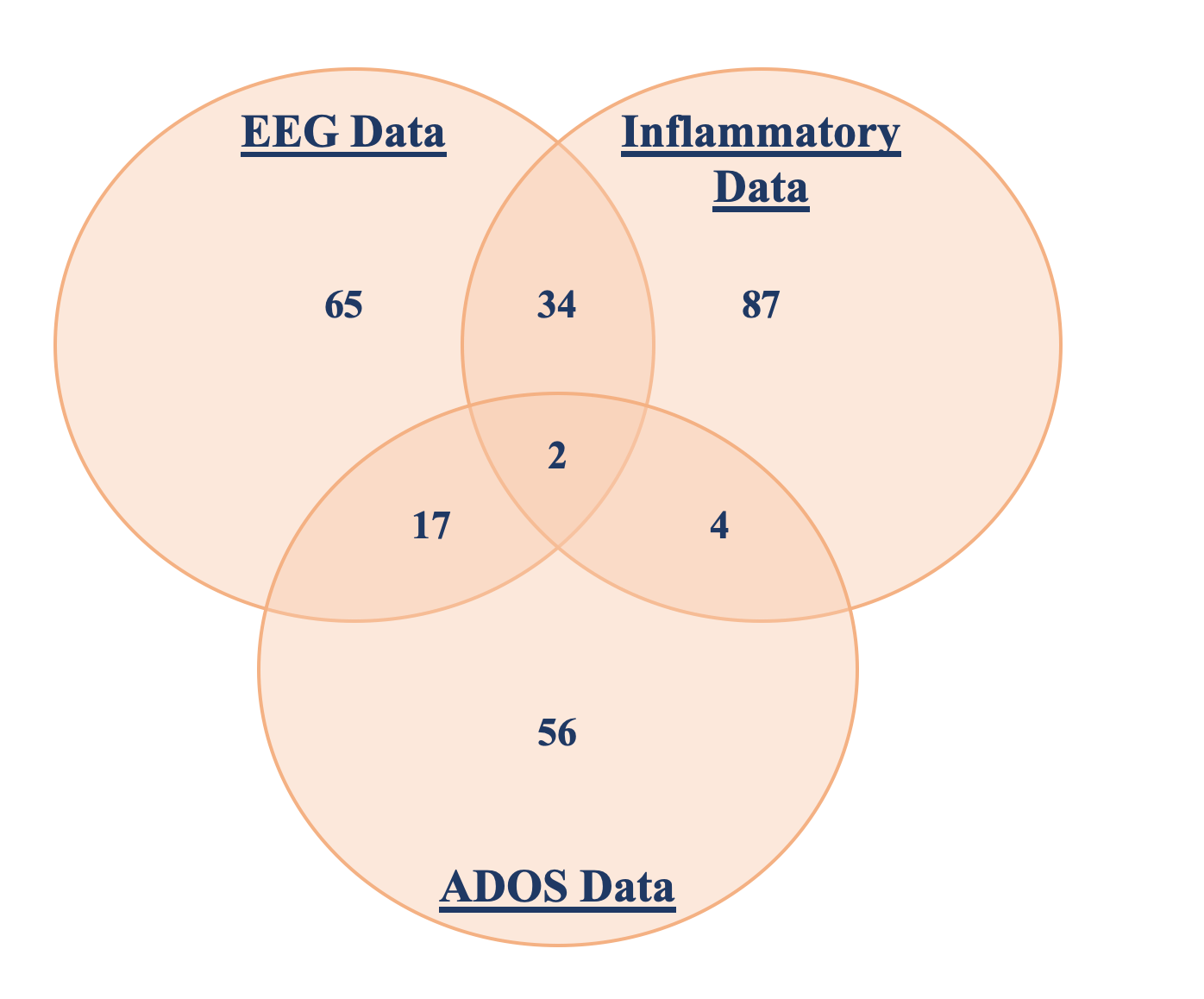

**Figure S1. Visualization of Data Overlap.** A visualization of the overlap in subjects between the three main types of data used in this study: EEG Data, Inflammatory Data, and ADOS data.

Figure S1 was created in order to better visualize which participants had usable data between and within the three major data categories: EEG Data, Inflammatory Data, and ADOS Data.

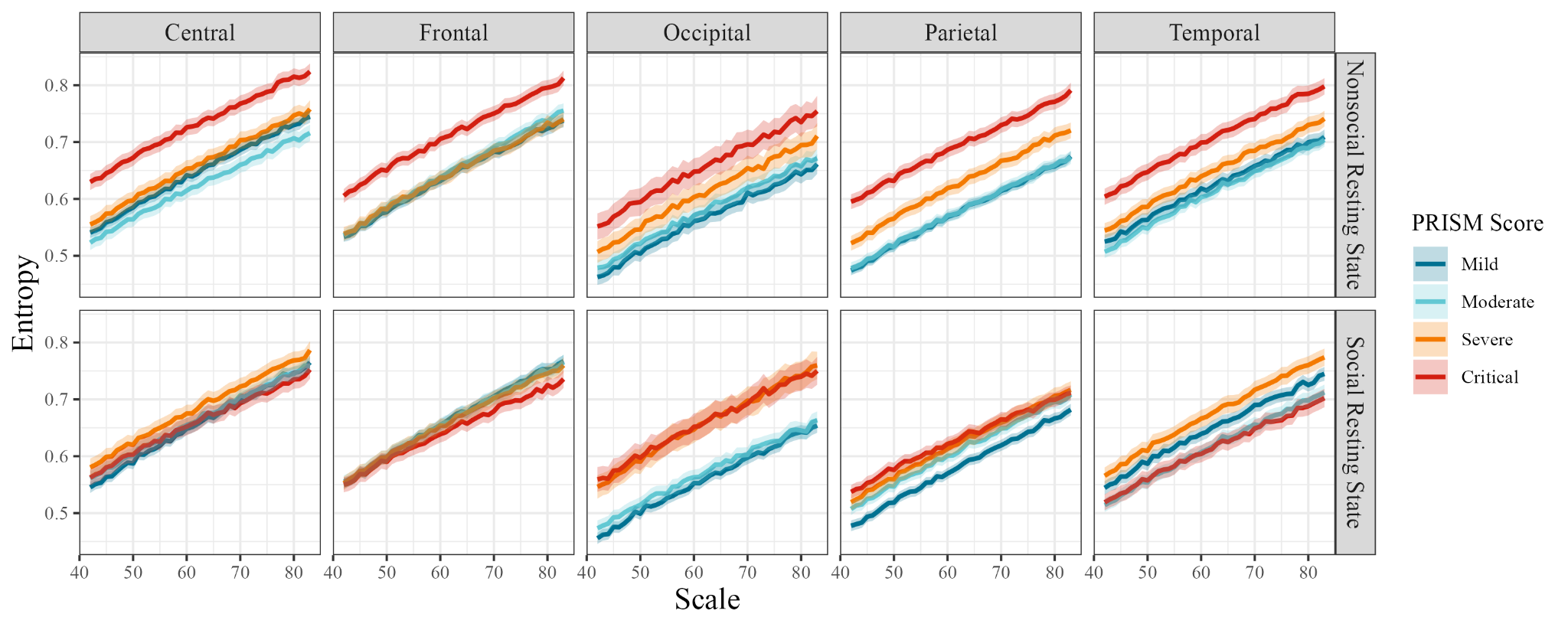

**Figure S2:** In the nonsocial resting state condition, across all brain regions, entropy is significantly different across all PRISM categories (Figure 2). In the social resting state condition statistically significant differences in entropy across PRISM categories differs depending on brain region.

We ran repeated measures ANOVA to determine the impact of brain region (Central, Frontal, Occipital, Parietal, Temporal) resting state condition (social vs. nonsocial), PRISM category, and scale on brain signal entropy (Figure 1). There was a significant three way interaction between resting state condition, PRISM category, and Brain Region. To further investigate these interactions we ran Tukey’s HSD tests within each combination of brain region and condition, the results of which can be seen in Table S1.

In the nonsocial resting state condition, across all brain regions, entropy is significantly different between the majority of PRISM categories (Figure S2). In the social resting state condition, differences in entropy across PRISM categories differ depending on brain region. In the occipital and temporal lobes, regions of the brain not typically associated with higher order social cognition^1^, entropy is significantly different across the majority of PRISM categories. However, in social brain regions like the central and frontal lobes, differences in entropy collapse. The central and frontal lobes function to integrate sensory information from the outside world^2^ and contribute to higher order cognition^3^, respectively. According to our analysis it appears that these regions of the brain, the seats of higher order cognition, are driving the overall lack of difference between PRISM categories observed in the whole brain, social resting state condition.

| **Effect** | **Degrees of Freedom** | **F statistic** | **P value** |
| --- | --- | --- | --- |
| Condition | 1.00 | 1.27 | 0.260 |
| PRISM Category | 3.00 | 400.05 | < 0.001 *** |
| Scale | 1.00 | 7699.18 | < 0.001 *** |
| Brain Region | 4.00 | 320.75 | < 0.001 *** |
| Condition x PRISM Category | 3.00 | 202.24 | < 0.001 *** |
| Condition x Scale | 1.00 | 0.01 | 0.910 |
| PRISM Category x Scale | 3.00 | 1.89 | 0.129 |
| Condition x Brain Region | 4.00 | 4.85 | < 0.001 *** |
| PRISM Category x Brain Region | 12.00 | 31.18 | < 0.001 *** |
| Scale x Brain Region | 4.00 | 0.76 | 0.553 |
| Condition x PRISM Category x Scale | 3.00 | 1.15 | 0.327 |
| Condition x PRISM Category x Brain Region | 12.00 | 18.63 | < 0.001 *** |
| Condition x Scale x Brain Region | 4.00 | 0.08 | 0.990 |
| PRISM Category x Scale x Brain Region | 12.00 | 0.42 | 0.958 |
| Condition x PRISM Category x Scale x Brain Region | 12.00 | 0.11 | 1.000 |
| **Comparison** | **Mean Difference** | **95% Confidence Interval** | **Adjusted P value** |
| **Central** | | | |
| **Nonsocial Resting State** | | | |
| Severe vs. Critical | -0.07 | [-0.09, -0.05] | < 0.001 *** |
| Moderate vs. Critical | -0.11 | [-0.13, -0.09] | < 0.001 *** |
| Mild vs. Critical | -0.08 | [-0.10, -0.06] | < 0.001 *** |
| Moderate vs. Severe | -0.04 | [-0.06, -0.02] | < 0.001 *** |
| Mild vs. Severe | -0.01 | [-0.03, 0.01] | 0.481 |
| Mild vs. Moderate | 0.02 | [0.002, 0.04] | 0.016 |
| **Social Resting State** | | | |
| Severe vs. Critical | 0.02 | [0.004, 0.05] | 0.006 ** |
| Moderate vs. Critical | 0.00 | [-0.02, 0.03] | 0.997 |
| Mild vs. Critical | 0.00 | [-0.02, 0.02] | 1.000 |
| Moderate vs. Severe | -0.02 | [-0.04, 0.0002] | 0.056 |
| Mild vs. Severe | -0.03 | [-0.05, -0.01] | < 0.001 *** |
| Mild vs. Moderate | -0.01 | [-0.03, 0.01] | 0.963 |
| **Frontal** | | | |
| **Nonsocial Resting State** | | | |
| Severe vs. Critical | -0.07 | [-0.09, -0.05] | < 0.001 *** |
| Moderate vs. Critical | -0.06 | [-0.08, -0.04] | < 0.001 *** |
| Mild vs. Critical | -0.07 | [-0.09, -0.05] | < 0.001 *** |
| Moderate vs. Severe | 0.01 | [-0.01, 0.02] | 0.992 |
| Mild vs. Severe | 0.00 | [-0.02, 0.02] | 1.000 |
| Mild vs. Moderate | -0.01 | [-0.03, 0.01] | 0.884 |
| **Social Resting State** | | | |
| Severe vs. Critical | 0.01 | [-0.005, 0.03] | 0.281 |
| Moderate vs. Critical | 0.02 | [-0.002, 0.04] | 0.139 |
| Mild vs. Critical | 0.02 | [-0.001, 0.04] | 0.085 |
| Moderate vs. Severe | 0.00 | [-0.02, 0.02] | 1.000 |
| Mild vs. Severe | 0.00 | [-0.02, 0.02] | 1.000 |
| Mild vs. Moderate | 0.00 | -1.83E-02 | 1.000 |
| **Occipital** | | | |
| **Nonsocial Resting State** | | | |
| Severe vs. Critical | -0.05 | [-0.07, -0.02] | < 0.001 *** |
| Moderate vs. Critical | -0.08 | [-0.10, -0.05] | < 0.001 *** |
| Mild vs. Critical | -0.09 | [-0.11, -0.07] | < 0.001 *** |
| Moderate vs. Severe | -0.03 | [-0.05, -0.01] | < 0.001 *** |
| Mild vs. Severe | -0.05 | [-0.07, -0.02] | < 0.001 *** |
| Mild vs. Moderate | -0.01 | [-0.04, 0.01] | 0.510 |
| **Social Resting State** | | | |
| Severe vs. Critical | 0.00 | [-0.02, 0.02] | 1.000 |
| Moderate vs. Critical | -0.09 | [-0.11, -0.07] | < 0.001 *** |
| Mild vs. Critical | -0.10 | [-0.12, -0.08] | < 0.001 *** |
| Moderate vs. Severe | -0.09 | [-0.11, -0.06] | < 0.001 *** |
| Mild vs. Severe | -0.10 | [-0.12, -0.08] | < 0.001 *** |
| Mild vs. Moderate | -0.01 | [-0.03, 0.01] | 0.678 |
| **Temporal** | | | |
| **Nonsocial Resting State** | | | |
| Severe vs. Critical | -0.06 | [-0.08, -0.04] | < 0.001 *** |
| Moderate vs. Critical | -0.10 | [-0.11, -0.08] | < 0.001 *** |
| Mild vs. Critical | -0.08 | [-0.10, -0.07] | < 0.001 *** |
| Moderate vs. Severe | -0.04 | [-0.06, -0.02] | < 0.001 *** |
| Mild vs. Severe | -0.03 | [-0.04, -0.01] | < 0.001 *** |
| Mild vs. Moderate | 0.01 | [-0.01, 0.03] | 0.614 |
| **Social Resting State** | | | |
| Severe vs. Critical | 0.06 | [0.04, 0.08] | < 0.001 *** |
| Moderate vs. Critical | 0.00 | [-0.02, 0.02] | 0.999 |
| Mild vs. Critical | 0.03 | [0.02, 0.05] | < 0.001 *** |
| Moderate vs. Severe | -0.06 | [-0.08, -0.04] | < 0.001 *** |
| Mild vs. Severe | -0.03 | [-0.05, -0.01] | < 0.001 *** |
| Mild vs. Moderate | 0.03 | [0.01, 0.05] | < 0.001 *** |
| **Parietal** | | | |
| **Nonsocial Resting State** | | | |
| Severe vs. Critical | -0.07 | [-0.09, -0.05] | < 0.001 *** |
| Moderate vs. Critical | -0.12 | [-0.14, -0.09] | < 0.001 *** |
| Mild vs. Critical | -0.12 | [-0.14, -0.10] | < 0.001 *** |
| Moderate vs. Severe | -0.05 | [-0.07, -0.03] | < 0.001 *** |
| Mild vs. Severe | -0.05 | [-0.07, -0.03] | < 0.001 *** |
| Mild vs. Moderate | 0.00 | [-0.02, 0.02] | 1.000 |
| **Social Resting State** | | | |
| Severe vs. Critical | -0.01 | [-0.03, 0.01] | 0.978 |
| Moderate vs. Critical | -0.02 | [-0.04, 0.002] | 0.109 |
| Mild vs. Critical | -0.05 | [-0.07, -0.03] | < 0.001 *** |
| Moderate vs. Severe | -0.01 | [-0.03, 0.01] | 0.641 |
| Mild vs. Severe | -0.04 | [-0.06, -0.02] | < 0.001 *** |
| Mild vs. Moderate | -0.03 | [-0.05, -0.01] | < 0.001 *** |

**Table S1 :** The results of the repeated measures ANOVA and post-hoc Tukey’s HSD investigating differences in entropy between PRISM categories across resting state conditions and brain regions.
